# Rates of Glaucoma Progression Derived from Linear Mixed Models Using Varied Random Effect Distributions

**DOI:** 10.1101/2021.06.01.21258173

**Authors:** Swarup S. Swaminathan, Samuel I. Berchuck, Alessandro A. Jammal, J. Sunil Rao, Felipe A. Medeiros

## Abstract

**Purpose:** To compare the ability of linear mixed models with different random effect distributions to estimate rates of visual field loss in glaucoma patients.

**Methods:** Eyes with ≥5 reliable standard automated perimetry (SAP) tests were identified from the Duke Glaucoma Registry. Mean deviation (MD) values from each visual field and associated timepoints were collected. These data were modeled using ordinary least square (OLS) regression and linear mixed models using the Gaussian, Student-t, or log-gamma (LG) distributions as the prior distribution for random effects. Model fit was compared using Watanabe-Akaike information criterion (WAIC). Simulated eyes of varying initial disease severity and rates of progression were created to assess the accuracy of each model in predicting the rate of change and likelihood of declaring progression.

**Results:** A total of 52,900 visual fields from 6,558 eyes of 3,981 subjects were included. Mean follow-up period was 8.7±4.0 years, with an average of 8.1±3.7 visual fields per eye. The LG model produced the lowest WAIC, demonstrating optimal model fit. In simulations, the LG model declared progression earlier than OLS (P<0.001) and had the greatest accuracy in predicted slopes (P<0.001). The Gaussian model significantly underestimated rates of progression among fast and catastrophic progressors.

**Conclusions:** Linear mixed models using the LG distribution outperformed conventional approaches for estimating rates of SAP MD loss in a population with glaucoma.

**Translational Relevance:** Use of the LG distribution in models estimating rates of change among glaucoma patients may improve their accuracy in rapidly identifying progressors at high risk for vision loss.

## Introduction

Detection of disease progression is essential in caring for patients with glaucoma. Standard automated perimetry (SAP) is the main testing modality used to evaluate functional vision loss in this patient population. An accurate assessment of rates of SAP change is essential in clinical decision-making to determine aggressiveness of therapy and follow-up. Identifying patients who exhibit fast rates of progression as soon as possible is paramount, as these individuals are at greatest risk for developing visual disability.

Estimation of rates of change has traditionally been made with ordinary least square (OLS) regression applied to global parameters such as mean deviation (MD) over time. However, OLS-derived estimates can be very imprecise in the presence of few measurements, a situation that is commonly seen in clinical practice. Previous studies have shown that, on average, clinicians end up obtaining less than one visual field test per year on glaucoma patients.^1^ With such low frequency of testing, OLS-derived rates of change would take more than 7 years to detect eyes progressing at moderate rate of visual field loss.^2^

OLS-derived rates of change utilize only measurements of the individual patient without accounting for the overall population from which the patient comes from. Previous work has shown that estimates of rates of change can be improved using linear mixed models,^3-5^ which allow data regarding the overall population to influence these estimates; the accuracy of estimates can be increased by “borrowing strength” from the population when fewer data points are available for a particular patient. Mixed model estimates include a fixed-effect component which represent the overall rate of a population, and a random-effect component that reflects the degree of deviation of an individual eye from the population average. This process creates eye-specific intercepts and slopes. Although not yet incorporated in routine clinical practice, estimates of rates of change using linear mixed models have been widely applied in research settings.^3-8^

A standard linear mixed model assumes that the random effects follow a Gaussian distribution. When applied to estimating rates of change this assumes that those rates are normally distributed in the population. However, it is known that only a relatively small proportion of glaucoma patients exhibit moderate or fast progression, which leads to a skewed distribution of rates of change in the population.^9-11^ Prior work has demonstrated that the assumption of normally distributed random effects may cause biased estimations of parameters when heterogeneity is present in a population, as would be expected in the rates of progression of glaucoma patients.^12^ Thus, fast progressors may not be properly identified due to shrinkage to the population mean in a Gaussian model.

Given how ubiquitous the use of mixed models is in glaucoma research and their potential for clinical applications, it is essential to determine whether the use of a normal distribution of random effects is appropriate in this context. In the present work, we investigated the impact of the random effects distribution on the estimates of rates of visual field loss and we assessed whether different distributions, such as Student t and log-gamma, would allow for more accurate estimation of rates of change and detection of eyes exhibiting fast progression.

## Methods

### Data Collection

The dataset used in this study was derived from the Duke Glaucoma Registry developed by the Vision, Imaging and Performance (VIP) Laboratory of Duke University.^13^ Institutional Review Board (IRB) approval was obtained for this analysis, and a waiver of informed consent was provided due to the retrospective nature of this work. All methods adhered to the tenets of Declaration of Helsinki for research involving human participants.

The database contained clinical information from baseline and follow-up visits, including patient diagnostic and procedure codes, medical history and imaging and functional tests. The study included patients previously diagnosed with primary open-angle glaucoma (POAG) or suspected of glaucoma based on International Classification of Diseases (ICD) codes. Patients were excluded if they presented with other ocular or systemic diseases that could affect the optic nerve or visual field (e.g. retinal detachment, retinal or malignant choroidal tumors, non-glaucomatous disorders of the optical nerve and visual pathways, atrophic and late-stage dry age-related macular degeneration, amblyopia, uveitis and/or venous or arterial retinal occlusion) according to ICD codes. Tests performed after treatment with panretinal photocoagulation (according to Current Procedural Terminology (CPT) codes) were excluded. ICD and CPT codes used to construct this database have been extensively detailed in a previous work.^13^ In addition, eyes that underwent trabeculectomy or aqueous shunt surgery were identified using CPT codes. For those eyes, only visual fields obtained before surgery were included, given the likely abrupt postsurgical alteration in the rate of change of SAP MD.

Glaucomatous eyes were identified as having an abnormal visual field at baseline (i.e., GHT “outside normal limits” or pattern standard deviation (PSD) probability <5%). Eyes suspected of glaucoma were identified with a “normal” or “borderline” GHT result or PSD probability >5% at baseline. All eligible subjects had SAP testing completed using the Humphrey Field Analyzer II or III (Carl Zeiss Meditec, Inc., Dublin, CA). SAP tests included 24-2 and 30-2 Swedish Interactive Threshold Algorithm (SITA) tests with size III white stimulus. Visual fields were excluded from this analysis if they had greater than 15% false-positive errors, greater than 33% fixation losses, greater than 33% false negative errors, or if the result of the glaucoma hemifield test (GHT) was “abnormally high sensitivity.” For this study, subjects were required to have ≥5 visual fields and ≥2 years of follow-up time.

### Model Formulation

OLS regressions were completed using standard linear regression for each eye. Bayesian linear mixed models were subsequently constructed. Bayesian statistics provide a probabilistic framework to address questions of uncertainty, such as the true rate of change in a glaucomatous eye. Prior distributions, which reflect an initial belief, are used in conjunction with available data (referred to as the likelihood) in order to generate estimates of specified parameters (posterior distributions.) For these models, a random-intercept and random-slope Bayesian hierarchical model was fitted for the SAP MD data:

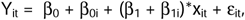

where Y_it_ represents the SAP MD value at time *t* of eye *i*, β_0_ represents the fixed intercept for the overall population, β_1_ represents the fixed slope for the overall population, and β_0i_ and β_1i_ represent eye-specific random intercepts and slopes respectively. In all models, the prior distributions for β_0_, β_1_, and the error term (ε_it_) were normally distributed. However, the prior distributions of the random effects differed as noted below. A correlation term with an unstructured correlation matrix was included in the model to account for associations between intercept and slope values. Of note, random effects were placed at the eye level; a more complex model with the eye nested within the patient did not provide additional improvement in the model, and thus the simpler model is described here. The correlation between intercept and slope was modeled using an unstructured covariance for the Gaussian and Student t models, while a covariance structure previously described was used for the log-gamma model.^14^

Gaussian, Student t, and log-gamma (LG) distributions were used to model the random effects of intercepts and slopes. The LG distribution is a left-skewed distribution that is sufficiently flexible to allow for more extreme negative values while maintaining a peak close to zero.^15^ Recent work has suggested that the LG distribution may be a more appropriate distribution in estimating the intercepts and slopes of MD and VFI, given the inherent left-skew of these data;^16^ the majority of eyes have values of intercepts and slopes near zero, but a smaller proportion of eyes have more extreme values.

In each model, the same distribution was used to model the random effects for both the intercept and slope. All statistical analyses were performed using R 3.6.3 (R Core Team, Vienna, Austria). For Gaussian and Student t distributions, the *brms* package in R was utilized. This package computes estimates of the posterior distributions using Stan, which is a C++ probabilistic Bayesian programming interface using Hamiltonian Monte Carlo (HMC) sampling (Stan Development Team, open source, 2018). HMC sampling is thought to be superior to traditional Markov chain Monte Carlo sampling, as this method can achieve a more effective exploration of the posterior probability space without inducing high rates of autocorrelation.^17^ For the LG distribution, the prior distribution was directly coded into Stan via the *rstan* R package.

### Data Analysis

Bayesian linear mixed models were compared using the Watanabe-Akaike information criterion (WAIC), a metric that reflects the overall fit of a Bayesian model. For each model, estimates of the posterior distributions of the parameters were obtained after running 4 chains with 8,000 iterations (burn-in of 1,000 iterations) per chain (i.e., a total of 28,000 iterations). These models were completed using high-performance computing servers on the University of Miami Triton supercomputer. Convergence of the generated samples was confirmed by evaluating trace plots and autocorrelation diagnostics. Summary measures, including posterior estimates of the fixed effects (β_0_ and β_1_), were calculated. Mean posterior estimated intercepts and slopes were calculated for each eye by adding the fixed and random effects of each draw and averaging these values for all draws corresponding to each eye. Eyes were defined as progressors if the one-sided Bayesian p-value was less than 0.05. OLS progressors were defined as those with a statistically significant negative rate of change (one sided p-value <0.05).

For predictive modeling, OLS and Bayesian models were constructed using different numbers of visual fields and assessing their ability to predict future observations. For example, a model using the MD values from the first 3 visual fields was constructed. This model was then used to generate a predicted value for the MD of the fourth, fifth, sixth, seventh, and eighth visual field. This process was repeated using the first 4 visual fields to predict the MD of the fifth, sixth, seventh, and eighth visual field and so on, up to a model that used the first 7 visual fields to predict the MD of the eighth visual field. The mean square prediction error (MSPE) of all Bayesian models and OLS were compared at each visual field. Bootstrapped 95% confidence intervals were calculated for MSPE for each model and at each visual field visit using 200 bootstrap samples. In addition to confidence intervals, we used analysis of variance (ANOVA) to perform a formal statistical hypothesis test that compared the MSPE across models. Tukey’s Honest Significant Difference test was used to test pairwise comparisons.

### Simulation Description

In preparation for simulations, the observed dataset from the Duke Glaucoma Registry was split in an 80%-20% fashion at the patient level. The 80% portion was used to train the Gaussian, Student t, and LG models that were subsequently used to evaluate the simulated eyes. The remaining 20% of the observed dataset was used to create a distribution of residuals for use in the simulations as detailed below.

In order to evaluate the ability of the models to estimate a diverse range of potential rates of change in glaucomatous and stable eyes, a set of simulated eyes was created. A total of 15 different “settings” were then generated from the combination of 3 intercept categories (mild, moderate, and severe) and 5 slope categories (non-progressor, slow, moderate, fast, and catastrophic). An intercept corresponding to mild, moderate, and severe disease at baseline was defined as an eye with a baseline MD between 0 and -6 dB, -6 and -12 dB, and -12 and -18 dB respectively. These values were chosen to simulate patients with mild, moderate, and severe glaucoma at baseline using the Hodapp-Anderson-Parrish classification system.^18^ Non-progressors were defined as those with a slope of 0 dB/year. Slow, moderate, fast, and catastrophic progressors were defined as eyes with a slope between 0 and -0.5 dB/year, -0.5 and -1.0 dB/year, -1.0 and -2.0 dB/year, and -2.0 and -4.0 dB/year respectively. These categories have been previously defined,^10, 13^ and were chosen to simulate eyes with varying rates of disease progression.

A total of 100 simulated eyes were generated for each setting, with the individual intercept and slope values randomly selected from the respective range of values. For each eye, a longitudinal sequence of visual field tests was simulated. Simulated timepoints of visual field testing were 0, 0.5, 1.5, 2.5, 3.5, 4.5, 5, 5.5 years. At each timepoint, the “true” MD value was based on the simulated intercept and slope. For example, assuming a “true” intercept of -4 dB and a “true” slope of -1 dB/year, “true” MD values would be -4, -4.5, -5.5, -6.5, -7.5, -8.5, -9, and -9.5 dB at the simulated timepoints. As visual field data are affected by noise, we added a residual value to each “true” MD value, according to a previously described methodology.^2, 19, 20^ As noted above, 20% of the observed data set was set aside and not used to train the models but rather was used to create a distribution of residuals binned to each dB value. This process constructs multiple distributions of residuals which reflect the heterogeneity in test variability that exists across the spectrum of disease severity. For each test in the sequence of visual fields, a residual value was randomly sampled from the distribution corresponding to the “true” MD. This noise component was then added to the “true” value. For example, for a “true” MD of -4 dB, the distribution of residuals corresponding to -4dB would be randomly sampled and a residual of +0.5 dB might be selected. This sampling would result in a simulated MD value of -3.5 dB. Using randomly selected residuals for the example above, a simulated set of values might be - 3.5, -3, -4.7, -7, -7.8, -9, -8.3, -11 dB. This simulated eye with these data points mimicking “real-world” observations and their inherent variability was then evaluated by OLS and Bayesian models as described below.

### Evaluating the Simulated Data

These 1,500 simulated eyes were then independently evaluated by the OLS, Gaussian, Student t, and LG models (which had been trained on the observed dataset) to obtain estimates of the eye-specific intercepts and slopes. Performance of the models was assessed within each simulation setting using bias and by calculating the rates of declared glaucomatous progression. Bias was defined as the difference between the true and estimated posterior slope; negative values of bias indicate underestimation of slope, positive values indicate overestimation of slope, and values closer to zero reflects more optimal prediction. Bias values were pooled across the intercept groups and were compared at each timepoint and for each progressor group using the Kruskal-Wallis test with the Dunn test for pairwise comparisons with Šidák correction. Given multiple comparisons, Bonferroni correction was applied, and an alpha value of 0.05/24 = 0.0021 was used to determine statistical significance. 95% credible intervals of bias for each model at each setting were also calculated. These intervals reflect that there is a 95% probability that the true value of bias lies within the calculated range. 95% credible intervals that exclude zero indicate significant underestimation of the slope (e.g., a 95% credible interval of (- 2.5, -0.5) indicates that the model significantly underestimates the slope.)

Rates of declaring progression were presented using cumulative event curves to compare the percentage of eyes that were declared to be progressing at each timepoint by the different models. To make these rates comparable, the p-value cutoff used for declaring progression in these simulated eyes was set to only allow 2.5% of non-progressor eyes to be erroneously identified as progressors (i.e., a false positive rate of 2.5%). P-value cutoffs were specified uniquely for each intercept range and time point. For each setting, the log-rank test was used to determine if the curves were significantly different. Finally, hazard ratios were calculated to determine if the Bayesian models differed in time to declaring progression, using Cox proportional hazards regression. Median time to declared progression was calculated for the different settings as the timepoint at which ≥50% of simulated eyes were declared as progressors.

## Results

The study included 6,558 eyes of 3,981 subjects with a mean age of 58.7±16.0 years at the time of the baseline visual field. A total of 52,900 visual fields were deemed reliable and evaluated. Mean follow-up period was 8.7±4.0 years, with an average of 8.1±3.7 visual fields per eye (range 5-34). **Table 1** contains additional patient characteristics. Female subjects comprised 58.2% of the cohort, while 31.5% identified as black. A total of 4,615 eyes (70.4%) had glaucomatous disease at baseline, while 1,943 eyes (29.6%) were suspected of glaucoma. Mean MD at baseline was -4.23±5.29 dB in the overall cohort. There was large variation in baseline MD of the eyes, ranging from -31.72dB to 2.58 dB.

**Table 1.** Demographics and clinical characteristics at baseline of the subjects included in the study.

| Characteristic | n = 6,558 eyes of<br>3,981 patients |
| --- | --- |
| Baseline age (years),<br>Mean ± SD | 58.7 ± 16.0 |
| Sex, female (%) | 2,356 (59.2) |
| Race, (%)<br>Black or African American | 1,254 (31.5) |
| **SAP** |  |
| Number of tests, n | 52,900 |
| Follow-up time (years),<br>Mean ± SD<br>Median (IQR) | 8.7 ± 4.0<br>8.1 (5.6; 11.2) |
| Number of tests per eye, n (%)<br>Mean ± SD<br>Median (IQR) | 8.1 ± 3.7<br>7.0 (6.0; 9.0) |
| Baseline SAP MD (dB),<br>Mean ± SD<br>Median (IQR) | -4.23 ± 5.29<br>-2.47 (-5.84; -0.71) |
| Baseline HAP glaucoma severity,<br>Mild, n (%)<br>Moderate, n (%)<br>Severe, n (%)<br>Glaucoma suspect, n (%) | 1,390 (21.2)<br>1,801 (27.5)<br>1,424 (21.7)<br>1,943 (29.6) |

*HAP = Hodapp-Anderson-Parrish; IQR = interquartile range; MD = mean deviation; SAP = standard automated perimetry; SD = standard deviation.*

Distributions of OLS slopes and the posterior estimated slopes of Bayesian models varied greatly (**Figure 1** & **Table 2**). The Gaussian model demonstrated a substantial shrinkage of estimates with a smaller range of slopes. In contrast, the range of slopes of the Student t model included both extreme negative and positive values, while the range of slopes of the LG model captured extreme negative values without extreme positive slopes (**Figure 2;** “eye-specific slopes” in **Table 2**). The LG model produced the lowest WAIC value, indicating that the LG model provided the optimal fit for the data compared to Gaussian and Student t models (**Table 2**). When comparing results of predictive modeling using a limited number of visual fields, Bayesian models consistently performed better compared to OLS, with lower MSPE values for each predicted visual field MD value (**Figure 2**). Overall mean MSPE values of the OLS, Gaussian, Student t, and LG predictions were 232.6±91.3, 5.2±0.3, 24.2±9.6, and 7.9±0.7 respectively, with significant differences noted between each Bayesian model and OLS (p<0.01 for pairwise comparisons) at each time point until 5 visual fields were utilized in the models. At this point, Student t predictions were no longer significantly different compared to those of OLS (p=0.84), but MSPE from the Gaussian and LG models remained significantly lower than those of OLS (p=0.01 and 0.02 respectively). Once 7 visual fields were utilized, all Bayesian model predictions were non-significant compared to OLS. Of note, differences in MSPE of the Gaussian, Student t, and LG predictions were not statistically significant at any time point.

**Figure 1.**
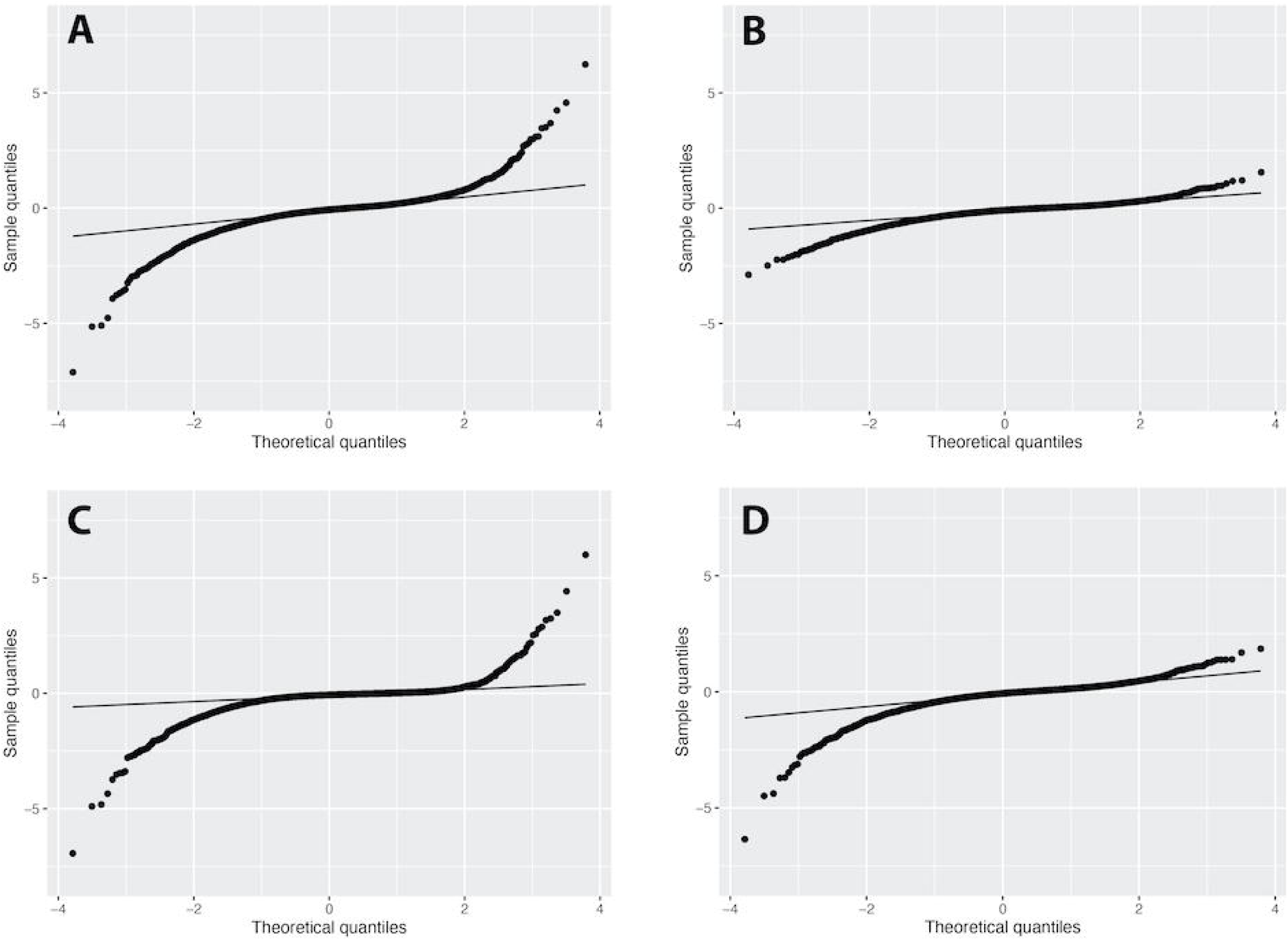
Quantile-quantile plots describing the distributions of the estimated slopes from (A) ordinary least square (OLS) regression and posterior estimated slopes from the (B) Gaussian, (C) Student t, and (D) log-gamma Bayesian linear mixed models. Deviations from the line indicate that the distributions are non-normal. OLS and Student t models demonstrated a wider range of slopes with more positive and negative extreme values, while the Gaussian model demonstrated a more normal distribution of slopes with shrinkage to the mean. The log-gamma model demonstrated more extreme negative values, indicating a left-skewed distribution.

**Figure 2.**
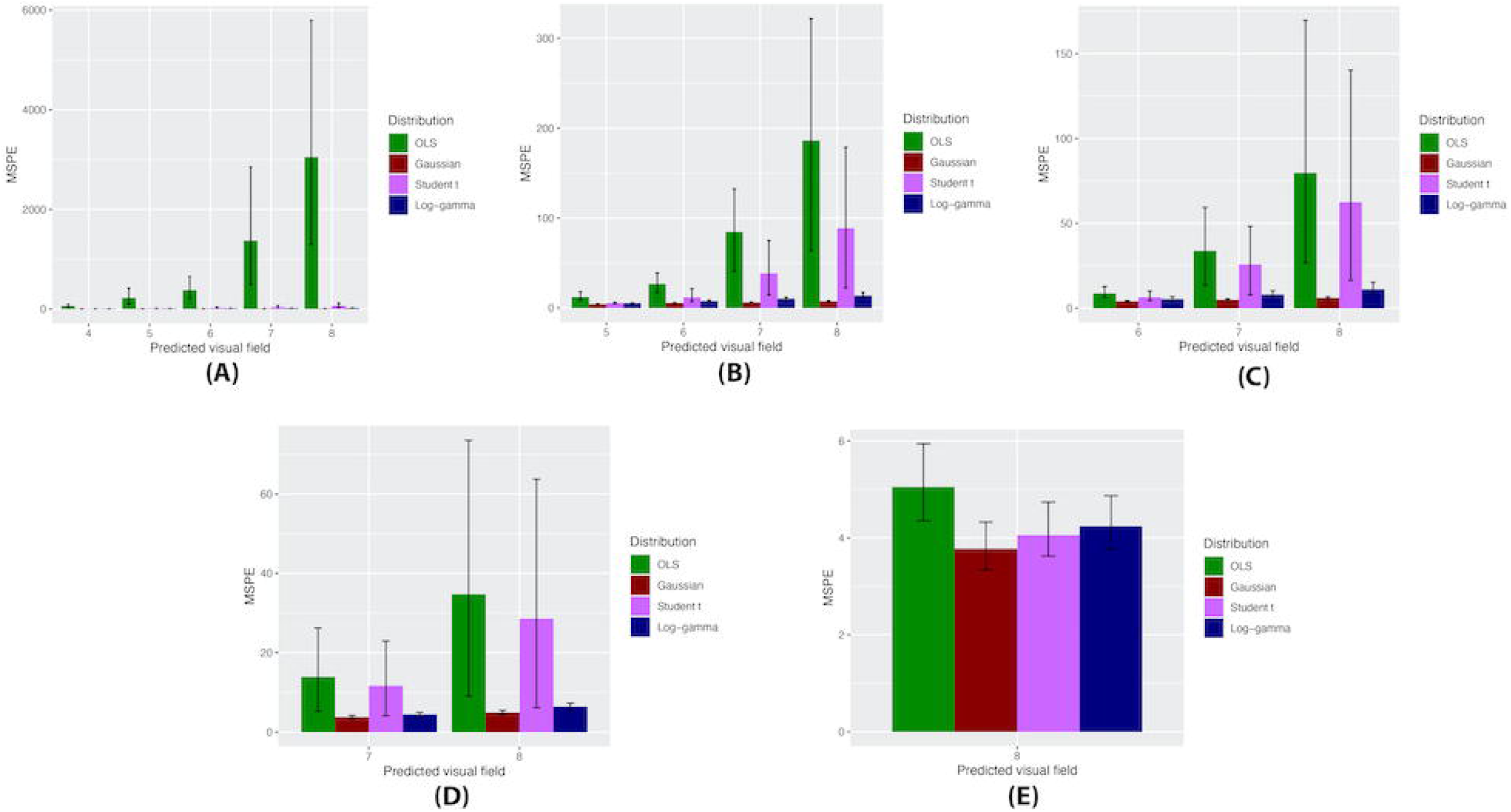
Mean squared prediction error (MSPE) of ordinary least square (OLS) regression and Bayesian linear mixed models in predicting the mean deviation of subsequent visual fields. MSPE values for the models constructed using the first (A) 3 visual fields, (B) 4 visual fields, (C) 5 visual fields, (D) 6 visual fields, and (E) 7 visual fields of an eye are shown. The x-axis represents the predicted visual field. Error bars represent bootstrapped 95% confidence intervals.

**Table 2.** Bayesian linear mixed model characteristics using varied random effect distributions. Eye-specific intercepts and slopes are estimates derived from the posterior distributions.

| | Population intercept ($\beta_0$) (dB) | Eye-specific intercepts (dB) | Population slope ($\beta_1$) (dB/year) | Eye-specific slopes (dB/year) | WAIC |
| --- | --- | --- | --- | --- | --- |
| Gaussian | -3.93 ± 0.06 | -2.24 (-5.37; -0.66) [-26.38; 5.07] | -0.15 ± 0.01 | -0.09 (-0.26; 0.01) [-2.89; 1.55] | 218,562 |
| Student t | -1.87 ± 0.04 | -2.15 (-5.15; -0.79) [-27.75; 5.87] | -0.07 ± 0.01 | -0.07 (-0.19; -0.01) [-6.93; 6.01] | 215,711 |
| Log-gamma | -3.80 ± 0.06 | -2.10 (-5.19; -0.66) [-27.47; 3.40] | -0.29 ± 0.01 | -0.07 (-0.29; 0.07) [-6.35; 1.86] | **214,188** |

Data displayed as mean ± standard error, or median (IQR) [range]
Boldface indicates lowest WAIC, demonstrating best model fit.
Eye-specific intercepts and slopes include both the fixed and random effect components.
*dB = decibel; WAIC = Watanabe-Akaike information criterion*

Distribution of slopes of all eyes and progressors from the various models are presented in **Tables 3** and **4** respectively. Compared to the Gaussian model, the LG and Student t models identified a greater number of eyes with faster rates of MD loss among all eyes and progressors. For example, the Gaussian model only identified 8.0% of all progressors as having fast progression and only 0.5% as having catastrophic progression. The LG model identified almost 2 times more progressor eyes as having fast progression (15.2%) and over 5 times more as having catastrophic progression (2.7%) (**Table 4**).

**Table 3.** Distribution of slopes estimated by ordinary least square (OLS) regression and Bayesian linear mixed models of all eyes. Slopes of the Bayesian models are estimates derived from the posterior distributions.

|  | < 0 to < -0.5<br>dB/year | < -0.5 to -1<br>dB/year | < -1 to -2<br>dB/year | < -2<br>dB/year |
| --- | --- | --- | --- | --- |
| OLS | 3,046<br>(46.4%) | 678<br>(10.3%) | 266<br>(4.1%) | 60<br>(0.9%) |
| Gaussian | 3,960<br>(60.4%) | 552<br>(8.4%) | 117<br>(1.8%) | 8<br>(0.1%) |
| Student t | 4,553<br>(69.4%) | 419<br>(6.4%) | 164<br>(2.5%) | 38<br>(0.6%) |
| Log-gamma | 3,181<br>(48.5%) | 642<br>(9.8%) | 222<br>(3.4%) | 39<br>(0.6%) |

Data presented as n (%) or mean ± standard deviation. *dB = decibels*

**Table 4.** Distribution of slopes estimated by ordinary least square (OLS) regression and
Bayesian linear mixed models of eyes identified as progressors. Slopes of the Bayesian models
are estimates derived from the posterior distributions.

|  | **Number of progressors** | **Mean Slope (dB/year)** | **< 0 to < -0.5 dB/year** | **< -0.5 to -1 dB/year** | **< -1 to -2 dB/year** | **< -2 dB/year** |
| --- | --- | --- | --- | --- | --- | --- |
| OLS | 1,679 | -0.62 ± 0.55 | 907<br>(54.0%) | 510<br>(30.4%) | 212<br>(12.6%) | 50<br>(3.0%) |
| Gaussian | 1,466 | -0.55 ± 0.32 | 792<br>(54.0%) | 549<br>(37.5%) | 117<br>(8.0%) | 8<br>(0.5%) |
| Student t | 1,452 | -0.58 ± 0.54 | 850<br>(58.5%) | 400<br>(27.5%) | 164<br>(11.3%) | 38<br>(2.6%) |
| Log-gamma | 1,463 | -0.69 ± 0.51 | 639<br>(43.7%) | 563<br>(38.4%) | 222<br>(15.2%) | 39<br>(2.7%) |

Data presented as n (%) or mean ± standard deviation. *dB = decibels*

Simulations demonstrated that the LG model was optimal in terms of accuracy as evidenced by the lowest degree of bias. Bias from the LG model was significantly lower than that of Gaussian and Student t models in all settings, most notably among fast and catastrophic progressors (**Figure 3**). Among fast and catastrophic progressors, mean bias from the LG, Student t, and Gaussian models was -0.51±0.49, -0.62±0.51, and -1.20±0.67 dB/year respectively (Kruskal-Wallis P=0.008). When evaluating 95% credible intervals of bias, Gaussian models persistently underestimated the true slope. Gaussian credible intervals excluded zero when using the first 3 visual fields among moderate progressors, when using the first 3, 4, or 5 visual fields among fast progressors, and when using the first 3, 4, 5, 6, or 7 visual fields among catastrophic progressors. In contrast, all 95% credible intervals of the Student t and LG models contained zero, indicating that these models did not severely underestimate the slope.

**Figure 3.**
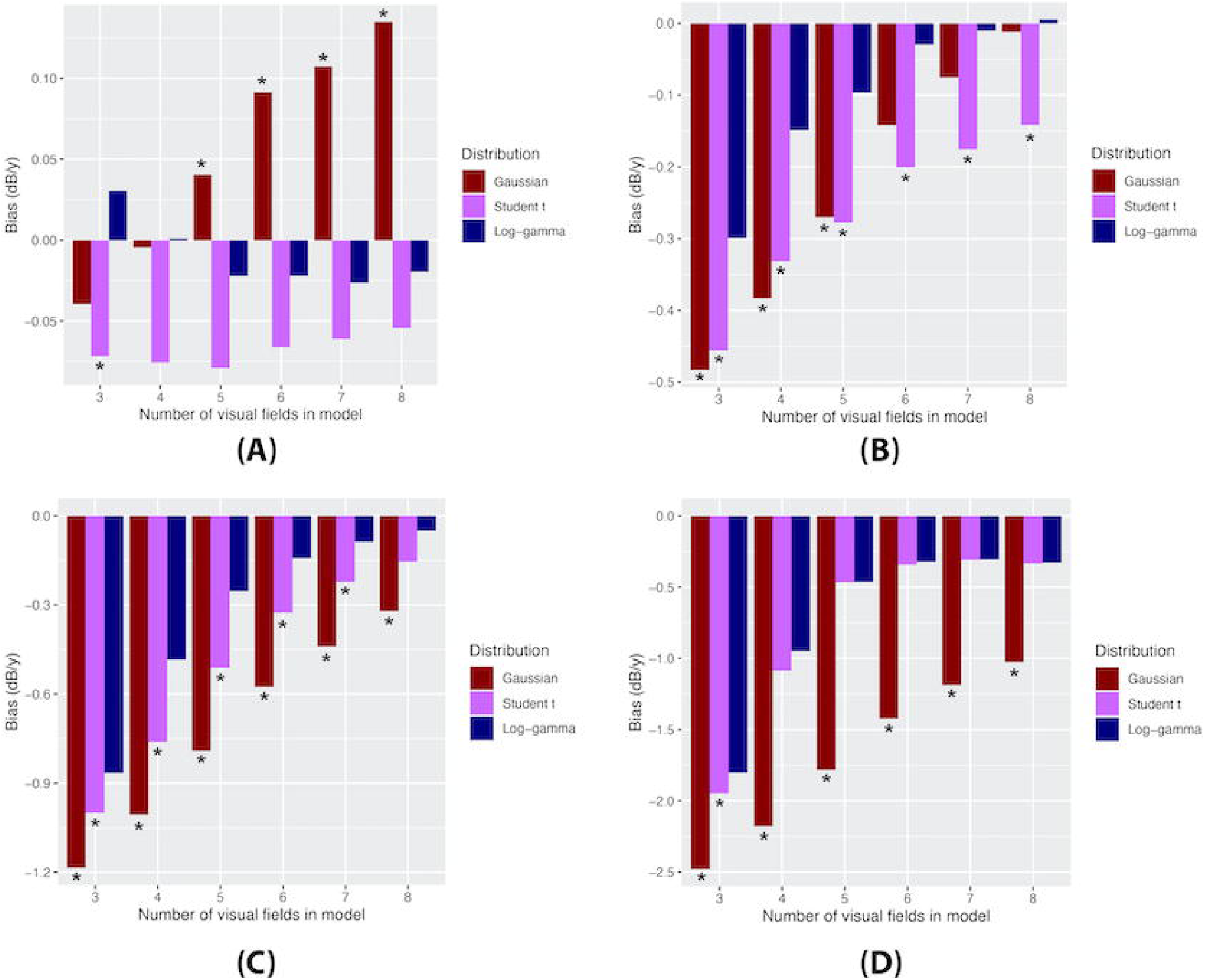
Comparison of bias in posterior estimated slopes of all Bayesian linear mixed models by number of visual fields used in the model to assess simulated eyes. Different progressor groups are shown: (A) slow, (B) moderate, (C) fast, and (D) catastrophic progressors. The asterisk (*) indicates a statistically significant difference between the LG model and the respective model using the Kruskal-Wallis and Dunn tests.

Cumulative event curves demonstrated a significant difference among the regression models (**Figure 4**; log-rank P <0.01). While all three Bayesian models performed similarly in terms of time to declaring progression (Cox hazard ratio P>0.05), they were significantly quicker to identify progression compared to OLS among moderate, fast, and catastrophic progressors (Cox hazard ratio P<0.001.) Median time to progression in the moderate, fast, and catastrophic progressors was consistently lower among Bayesian models compared to OLS (**Table 5**). The average median time to progression was lower in the LG model (2.8 years) compared to Student t (3.0 years), Gaussian (3.2 years), and OLS (4 years).

**Figure 4.**
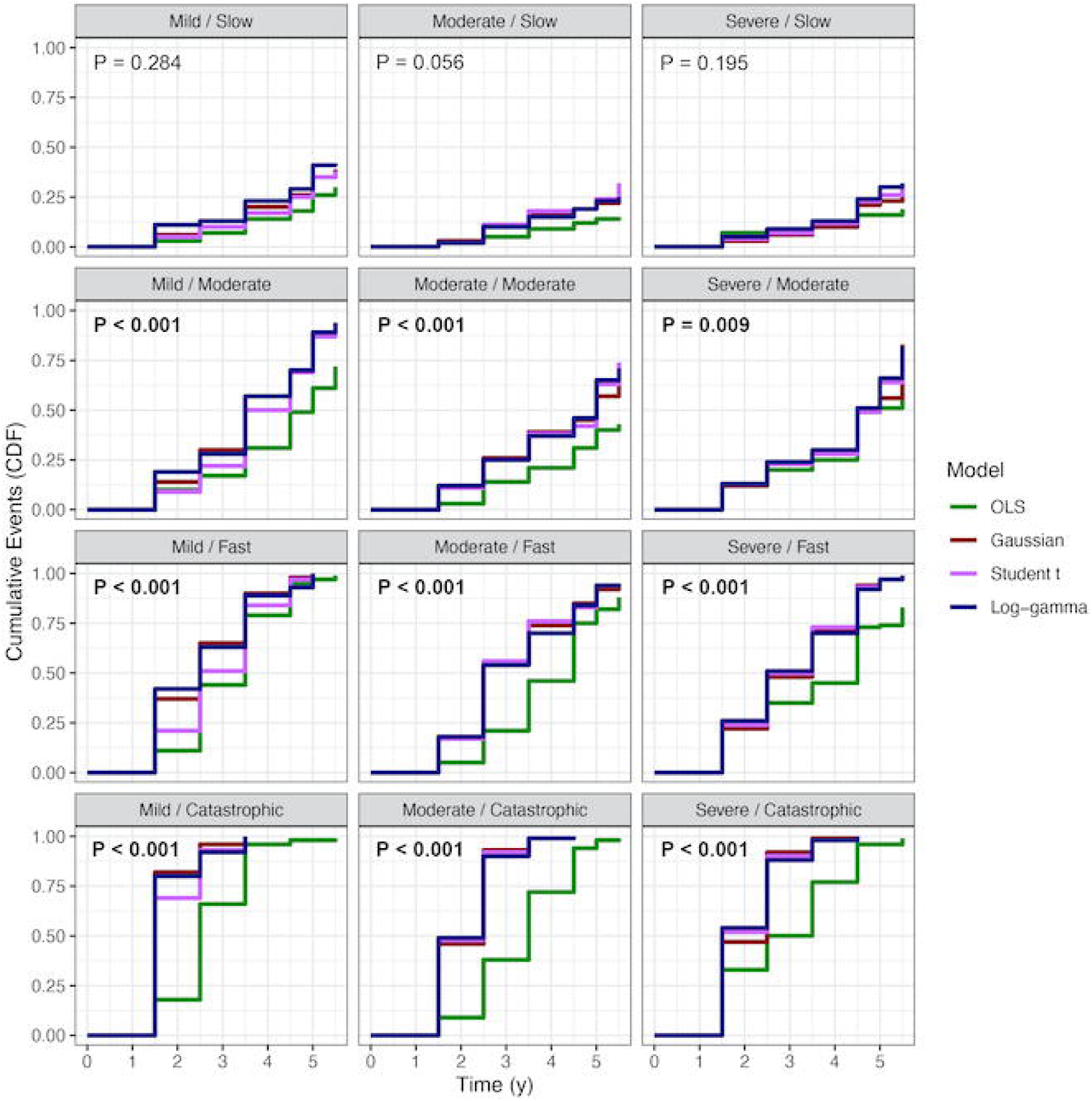
Cumulative event curves demonstrating cumulative probability of declaring glaucomatous progression in various simulation settings. The first term of each setting refers to the baseline disease severity, while the second refers to the rate of change. The curves of the three Bayesian linear mixed models and ordinary least squares (OLS) regression were compared with the log-rank test, and the respective p-values are presented for each setting. Of note, the third visual field occurred at 1.5 years and the fifth visual field occurred at 3.5 years in these simulations.

**Table 5.** Median time to declared progression (in years) among different progressor groups with
ordinary least squares (OLS) regression and Bayesian linear mixed models. None of the models
reached 50° in declared progression after 5.5 years for slow progressors. The third, fourth,
fifth, sixth, and seventh visual fields occurred at 1.5, 2.5, 3.5, 4.5, and 5.0 years respectively.

|  | **Slow** | **Moderate** | **Fast** | **Catastrophic** |
| --- | --- | --- | --- | --- |
| OLS | N/A | 5.0 | 4.2 | 3.2 |
| Gaussian | N/A | 4.5 | 2.8 | 2.2 |
| Student t | N/A | 4.8 | 2.5 | 1.8 |
| Log-gamma | N/A | 4.3 | 2.5 | 1.8 |

## Discussion

In this study, we compared the effect of various random effect distributions on estimating rates of visual field change using Bayesian linear mixed models with a large dataset of over 6,000 eyes. Bayesian models provided significantly improved predictions compared to conventional OLS regression when only a limited number of visual fields were available. Among the distributions tested for Bayesian models, the LG was optimal in terms of overall model fit with the lowest WAIC value. In addition, simulations showed that the LG model had the lowest bias and was sufficiently flexible to rapidly identify fast progressors. These findings suggest that Bayesian models using the LG distribution may offer significant advantages compared to more traditional approaches in modeling rates of change in glaucoma.

Our results showed the value of the Bayesian models compared to OLS regression when estimating rates of change in the presence of relatively few observations. Bayesian models consistently outperformed OLS in quickly declaring progression, especially among fast and catastrophic progressors. For example, after only 1.5 years (3 visual fields) in the “mild / catastrophic” setting **(Figure 4**), LG and Gaussian models declared progression in over 80% of true progressors, while OLS only detected 18% of progressors. Wu et al previously demonstrated that using OLS, 80% of eyes progressing at -2 dB/year would be identified as progressors only after 2.1 years if 3 visual fields were performed per year (i.e., after 6 visual fields were completed).^2^ While the benefit of Bayesian linear mixed models over OLS appeared to decline once 7 tests were available (**Figure 2**), obtaining visual fields at a sufficiently high frequency to procure such a large number of tests is often challenging in clinical practice. The reduction in time to progression using a minimal number of visual fields with Bayesian modeling may be of great value to the clinician. Median time to progression was lower among Bayesian models, especially with the LG model (**Table 5**).

The LG model demonstrated the greatest accuracy with the lowest amount of bias among different progressor groups (**Figure 3**). Zhang et al previously demonstrated the value of the LG model, as it provided a better fit for SAP data derived from 203 patients in a prospective study compared to a Gaussian model.^16^ The authors also constructed a joint longitudinal model using functional SAP and structural optical coherence tomography data, which demonstrated a stronger correlation between functional and structural rates of change when the LG model was utilized. Our work confirms the better fit of the LG model in a much larger dataset.

We found it interesting that the Bayesian models identified fewer eyes as progressors compared to OLS. Prior studies had indicated that OLS identified fewer progressors compared to Bayesian models, although these studies evaluated smaller datasets of eyes with fewer number of tests.^3, 4^ We believe that this discrepancy is due to the greater number of tests that were available in the current dataset (mean of 8.1 visual fields per eye). When evaluating those eyes identified as progressors by OLS but not by the Gaussian, Student t, and LG models, the average OLS rate of change was -0.30±0.25 dB/y (IQR -0.35 to -0.14), -0.31±0.26 dB/y (IQR - 0.38 to -0.14), and -0.28±0.21 dB/y (IQR -0.34 to -0.14) respectively. These values are reflective of slow rates of change, which would not be as worrisome to the clinician and would be unlikely to lead to severe vision loss. In contrast, when evaluating eyes identified as progressors by Gaussian, Student t, and LG models but not by OLS, the average OLS rate of change was - 0.64±0.57 dB/y (IQR -0.78 to -0.30), -0.62±0.57 dB/y (IQR -0.76 to -0.26), and -0.68±0.58 dB/y (IQR -0.86 to -0.31) respectively. OLS was unable to confirm progression among these eyes displaying a faster rate of change, which would be of greater concern and clinical importance. Although the Bayesian models may have identified fewer progressors, the clinical relevance of the progressors identified by these models appears to be greater.

Given the higher percentage of eyes with faster rates of change in the LG model (**Table 4**), one might also be concerned about overestimation of slopes. However, bias data from the simulations demonstrated that 95% credible intervals of the LG model never included 0, indicating that this model did not significantly overestimate rates of change. Although MSPE values were comparable between LG and Gaussian models **(Figure 2**), the LG model was able to estimate the rate of fast and catastrophic progressors more accurately. In contrast, the Gaussian model underestimated these rates, with bias values twice as large on average. In the observed data, the Gaussian model was more likely to shrink estimates closer to the population mean (**Tables 3** and **4**). These findings serve as a warning that linear mixed models using the Gaussian distribution to describe visual field data will likely underestimate the rates of change among this subset of patients. These individuals are arguably the most important to identify since they are at high risk for visual disability. Although most glaucoma patients will progress if followed for a sufficient amount of time, rates of change vary greatly.^3^ The magnitudes of these rates are crucial to clinical care; while slow progressors may be carefully observed, fast progressors may need to be treated more aggressively in order to prevent vision loss. Therefore, accurate estimation of rates of change is essential to characterizing the nature of a patient’s disease. The LG model was able to accurately identify fast progressors while still characterizing the majority of eyes as slow or non-progressors.

Limitations of this study include the assumption that eyes exhibit a linear rate of change over time. Although it is likely that visual field losses are non-linear over the full course of the disease,^21-23^ a linear approximation is likely a reasonable approximation within the limited timeframe used to make most clinical decisions. We also assumed a constant correlation between intercept and slope regardless of disease severity. In clinical practice, severe glaucoma patients are often aggressively monitored and treated, leading to a reduction in correlation between baseline disease (i.e., the intercept) and rate of change (the slope). In addition, the retrospective data collection does not provide insight into augmentation of medical therapy, which could affect rate of progression. It is possible that additional medical or laser therapies may have occurred between visual field tests. The censoring protocol described above only pertained to surgical glaucoma cases. Finally, other potential distributions exist to model random effects which were not evaluated in the present study. We empirically chose Gaussian, Student t, and LG for comparison given clinical knowledge regarding the distribution of SAP MD in large populations. Further studies should investigate whether other distributions may provide advantages compared to the ones assessed in our work.

In summary, we have demonstrated that a Bayesian hierarchical model using the LG distribution provides the optimal model fit for a large SAP dataset compared to Gaussian and Student t distributions. The LG model is sufficiently flexible to accurately characterize non-progressors, slow progressors, and fast progressors. While Gaussian and LG models are comparable in predicting future SAP MD values, Gaussian models tend to underestimate fast progressors. The LG model was optimal in predicting the rates of change with greatest accuracy while rapidly identifying progressors. These findings may have significant implications for estimation of rates of visual field progression in research and clinical practice.

## Data Availability

Data is available upon reasonable request.

